# A data first approach to modelling Covid-19

**DOI:** 10.1101/2020.05.22.20110171

**Authors:** Jayanti Prasad

**Affiliations:** Khagol-20, 38/1, Panchavti, Pashan, Pune (India) - 411008

## Abstract

The primary data for Covid-19 pandemic is in the form of time series for the number of confirmed, recovered and dead cases. This data is updated every day and is available for most countries from multiple sources such as [Gar20b, iD20]. In this work we present a two step procedure for model fitting to Covid-19 data. In the first step, time dependent transmission coefficients are constructed directly from the data and, in the second step, measures of those (minimum, maximum, mean, median etc.,) are used to set priors for fitting models to data. We call this approach a “data driven approach” or “data first approach”. This scheme is complementary to Bayesian approach and can be used with or without that for parameter estimation. We use the procedure to fit a set of SIR and SIRD models, with time dependent contact rate, to Covid-19 data for a set of most affected countries. We find that SIR and SIRD models with constant transmission coefficients cannot fit Covid-19 data for most countries (mainly because social distancing, lockdown etc., make those time dependent). We find that any time dependent contact rate decaying with time can help to fit SIR and SIRD models for most of the countries. We also present constraints on transmission coefficients and basic reproduction number 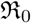, as well as effective reproduction number 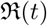. The main contributions of our work are as follows. (1) presenting a two step procedure for model fitting to Covid-19 data (2) constraining transmission coefficients as well as 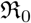 and 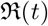, for a set of countries and (3) releasing a python package **PyCov19** [Pra20b] that can used to fit a class of compartmental models, with time varying coefficients, to Covid-19 data.

## 1 Introduction

At present the world is going through an unprecedented crisis of pandemic Covid-19 caused by a novel form of coronavirus, named Sars-CoV-2 which was passed to the human from bats in the Wuhan city of China, some time in December 2019 [Org20a, Org20b, ea20h, ea20t, ea20v, ea20d, ea20r, ea20l]. Till the end of May 2020 the virus has reached almost all the parts of the world resulting in more than six million people infected and more than a quarter million deaths [Wor20]. The measures to contain the virus medically by developing a vaccine are going on war footing. However, the success is still expected to be a few years away [ea20f]. Till a fraction of the population develop (herd) immunity or the vaccine is ready, the only means to contain the pandemic are social measures (social distancing, contact tracing etc.,) and enhanced hygiene practices [ea06, ea20s, ea20p].

Some of the most important problems related to Covid-19 research are (1) estimating the controlling parameters of the pandemic, (2) making short term predictions using mathematical-statistical modeling which can help in mitigating policies (3) simulating the growth of the epidemic by taking into account as many contributing effects as possible and (4) quantifying the impact of mitigation measures, such as lockdown etc [ea20j].

Modeling Covid-19 pandemic with compartmental models of Kermack and McKendrick (for an introduction see [JR08, Li18, BC18]) has been one of the most active problems in the recent times [ea20p, ea20a, ea20c, ea20e, FP20, ea20m, Oli20]. There have been alternative approaches also such as [IM20] where statistical considerations are being taken into account for predictions. In one of the studies [FP20] it is argued that the data for the confirmed, recovered and dead, all three can easily fit a power law model with similar coefficients. The main attractive feature of these data driven approaches is that the complexity of the model being considered is determined by the data and not by theoretical expectations.

In the present work we follow a middle approach and fit two compartmental models, named SIR and SIRD with some modification, to the Covid-19 data. One of the main reasons to consider these models has been that the Covid-19 data is available only for the **S**usceptible, **I**nfected, **R**ecovered and **D**ead compartments (for the notations used here and other places in the present work see table (1)). It may be true that a large fraction of the population which may be **E**xposed (defined later) play an important role in the dynamics of the pandemic however, it is hard to get reliable numbers for that. Apart from that, a large number of undocumented cases [ea20l] may also have significant influence on the spread of the pandemic.

**Table 1:**
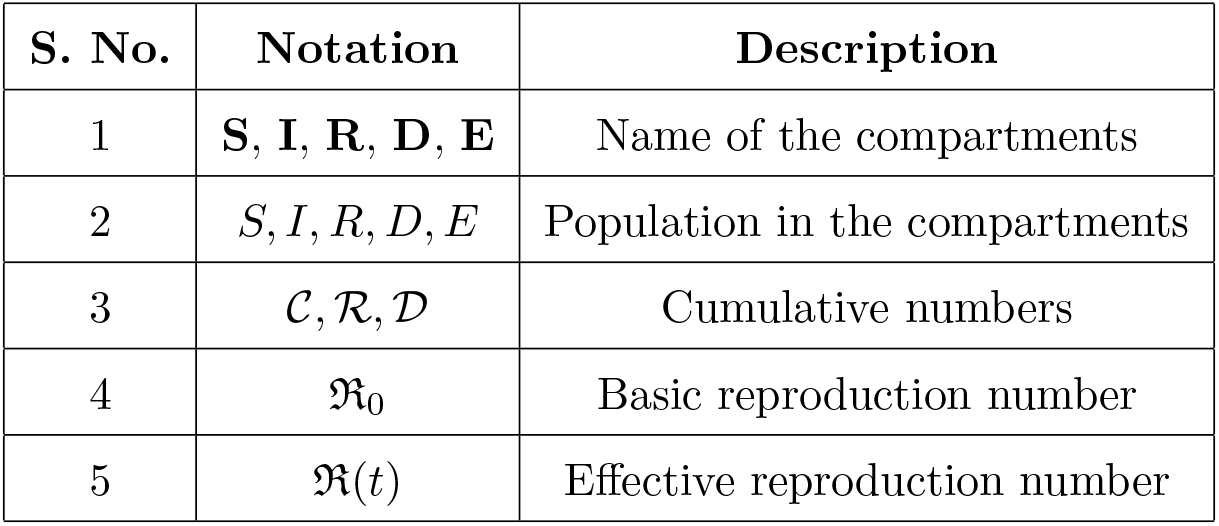
Notations used in the work

A brief summary of the work presented here is as follows.

In §2 we give a brief introduction to the compartmental models and introduce the notations and variables used in the work. In particular, we discuss the SIR model in §2.1 and the SEIR and the SIRD models in §2.2 and §2.3 respectively. One of the major parts of the work presented here is to study the time dependence of the contact rate *β*, we introduce a set of parametric models of *β*(*t*) in §2.4. We discuss the time series data used in the study in the §3 by giving an example of Italy which is one of the most affected countries. The main results of our work are given in §4 and in §5. In §4 we discuss the reconstruction (regression) procedure for the set of transmission coefficients as well as for the effective reproduction (defined later), number 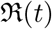. Parameter estimation is discussed in §5. The main conclusions of our work with a summary and some important points are discussed in §6.

## 2 Mathematical Modeling

Mathematical modeling of epidemics that started a century back with the seminal work of Kermack and McKendrick (see [NC08, JR08, Li18] for an introduction) is still the main framework most commonly used. The main idea of the Kermack and McKendrick’s compartmental models is that every individual in a society belongs to one of the *M* compartments and the total number of individuals belonging to different compartments keep changing with time. The minimum value *M* can have is two, for the Susceptible-Infected-Susceptible (SIS) model, in which the recovery does not guarantee that one will not get the infection again [JR08].

During an epidemic phase an individual can go through many stages from being perfectly healthy to the recovered one after an infection, with or without any immunity (short or long term) or may die. If we represent every stage with a compartment and keep the track of the number of individuals in each compartment then we can easily model the dynamics of the epidemic. This approach is very similar to the approach taken in astronomy where we count the number of stars in different stages of their life to understand the stellar evolution.

In principle we can have any number of logical compartments but in practice we should consider only those compartments for which we have the counts data, in particular for model fitting. Taking into account the fact that we have data only for the number of confirmed, recovered and dead population, the only compartmental model that meets the requirement is the SIRD model. If we consider the recovered and dead together we get the SIR model as is discussed in the next section.

One of the important compartments that also is commonly considered is the ‘exposed’ one and represents the population which have received the infection but cannot pass to others, before a certain period called the incubation period. If we consider exposed population also then we get the SEIR model that also is discussed below. Three compartmental models SIR, SIRD, and SEIR are shown in the (a), (b) and (c) panels of Figure (1) respectively (for more detail one can refer to [JR08, Het00, ea97, Oli20]).

**Figure 1:**
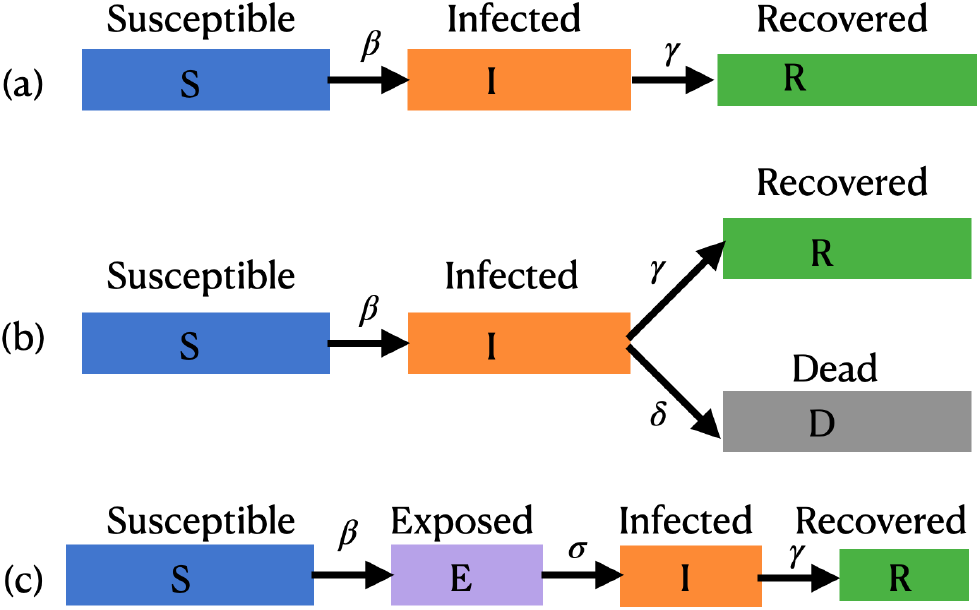
In the compartmental models the total population *N* is divided into a set of compartments as shown in the figure. The number of compartments and their connectivity depends on the detail of the model being considered. If we consider these compartments as nodes of a graph then there are transmission coefficients for every connecting edge that determine how effective that edge is in changing the population of the connected compartments. In (a) and (c) representing SIR and SEIR models, the compartments are connected in a linear way, however, for the case (b), representing the SIRD model, there is a branching also. Since the total population must remain a constant so the rates of change along all the connecting edges must add to zero.

If we identify the compartments with the nodes of a graph then the transmission between different compartments, as is represented by a set of coefficients, can be considered the edges of the graph. Some of the nodes may have multiple edges and some of the edges could be bi-directional also. The main challenge of the modeling a pandemic like Covid-19 is not the scarcity of mathematical models but it is of the reliable data for the compartments being considered.

### 2.1 SIR Model

The most basic compartmental model is the SIR model which is shown in (a) of the Figure (1) and is described by the following set of equations:

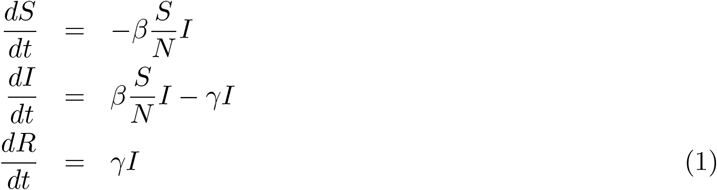

Here *β* and *γ* are the transmission coefficients, also called the contact rate and the recovery removal rate respectively, and 1*/β* and 1*/γ* represent the mean duration of infectiousness and the average period of infectivity (see [Het00, ea03a]), respectively, (see Table (1 for notations).

In general there is some time lag between acquiring an infection and becoming infectious. However, in the SIR model it is ignored and an assumption is made that individuals become infectious immediately upon getting an infection. This is a very strong assumption and the main reasons for making this is that we do not know reliably how many people are actually ‘exposed’, or have the virus but are still not infectious (cannot pass it to others). One of the ways to address this problem could be contact tracing and assuming that anyone who has come into contact with an infected person is an exposed one. However, this assumption is as strong as the assumption made in the SIR model.

If we do not consider the birth, death and movement of people then the following condition must be satisfied.

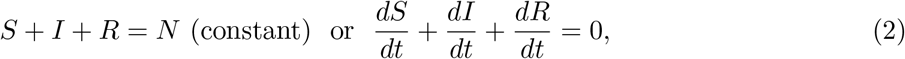

here *S, I* and *R* is the population of the **S**, **I** and **R** compartments respectively.

In equation (1) the transmission coefficient *β* is one of the most important parameters of the epidemic dynamics and can be written as the product of the contact rates (the average number of contacts per person per time) and the transmission probability (the probability of disease transmission on contact between a susceptible and an infectious person). As has been mentioned that the transmission coefficient *γ* can be identified with the recovery rate which is nothing but the inverse of the infectious period (during which an infected person can pass the virus to other healthy people).

In general, the equations (1) is solved with the following initial conditions:

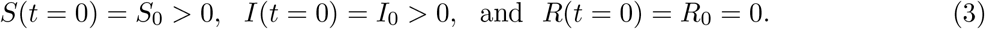

The second equation from 1 can be written as:

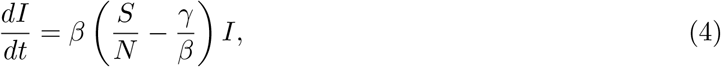

and for *S/N > γ/β* we get a positive infection rate.

Here we define one of the most important parameters of an epidemic in terms of the ratio *β/γ*, called the **basic reproduction number** 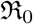, when considered a constant, and called the **effective reproduction number** 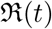, when considered a function of time. The most common definition [JR08] of it is that it is *the average number of secondary cases arising from an average primary case in an entirely susceptible population*. Note that in the text we may also use just “reproduction number” and the meaning of it will depend on the context. Some of the studies such as [ea20a] call 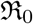 and 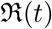 both as basic reproduction number, however, we follow the convention used in [ea03b, ea03a, Cob20].

The basis reproduction number 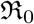 is the main measure which quantifies the transmissibility of the virus and 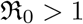, sets a chain of transmissions leading an exponential growth of the pandemic. We can keep 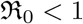, by minimizing the contact rates (social distancing etc.,), lowering the infectiousness of the infected people (by treating them or putting them in a quarantine etc.) and reducing the susceptibility of the healthy people by vaccination etc., (for detail see [ea05]).

The SIR model is one of the most basic models and can be easily generalized by one or more of the following ways:

1. **Adding more compartments:** Depending on the type of pandemic and other details we are interested in we can add more compartments to the SIR model. These compartments can fit in between the existing ones (for example as shown in Figure (1) (c) for SEIR case) or can branch out from the existing once (as shown in Figure (1) (b) for the SIRD case). With every new compartment added we must include the transmission coefficients for the connecting edges and also need an initial population for the new compartment being added [BC18, BS20, ea20b, ea20u].
2. **Heterogeneous population:** In the basic SIR model we consider a homogeneous population which share transmission coefficients such as *β* and *γ*. This may not be true in practice, for example, people from different groups (based on age, medical conditions, gender etc, etc) may have different contact rate *β* and/or recovery rate *γ* [ea20p, RR20].
3. **Variable transmission coefficients:** Theoretically transmission coefficients such as *β* and *γ* are considered constant, however, in practice they can vary with time due to multiple factors as the pandemic spreads [Cob20, P.20, GPD20]. For example, social distancing and other precautions such as hand wash etc., may help to lower *β*. In a similar way a better understanding of the disease and the urgency with which medical and testing facilities are brought online may improve the recovery rate *γ*.

**Figure 2:**
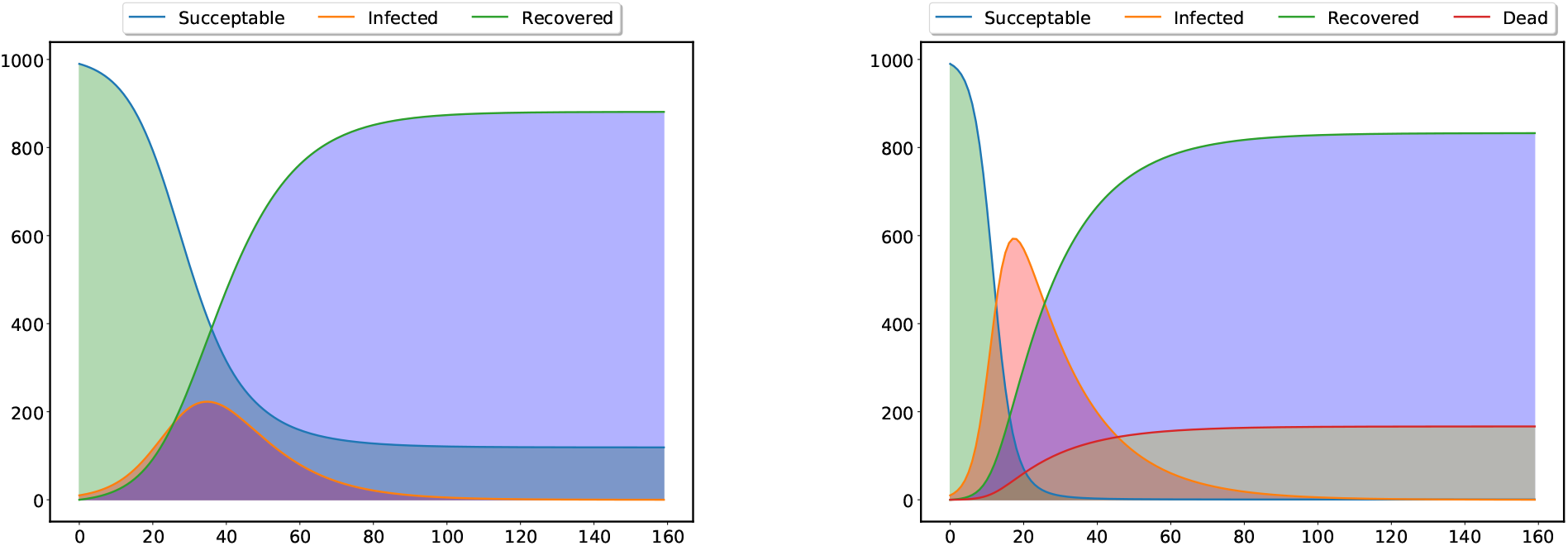
The left panel in the figure shows the standard SIR model with three type of population (compartments) - Susceptible, Infected and Recovered and the right panel shows the SIRD model with four type of population - on extra compartment for the dead population. The dynamics of the epidemic depends on the transmission coefficients *β*, *γ* and *δ* and the starting values of the different types of population. The values of the parameters and the initial population in different compartments used in the figure are only for the purpose on an illustration.

### 2.2 SEIR model

If we relax the assumption that the people who get the infection become infectious instantly and consider a latent period to the the onset of infectiousness there is a fraction of population (compartment) which has been exposed to the virus but will become infectious only after some latent period 1*/σ*, then the model is called **S**usceptible-**E**xposed-**I**nfected-**R**ecovered (SEIR) model represented by the following set of equations [JR08]

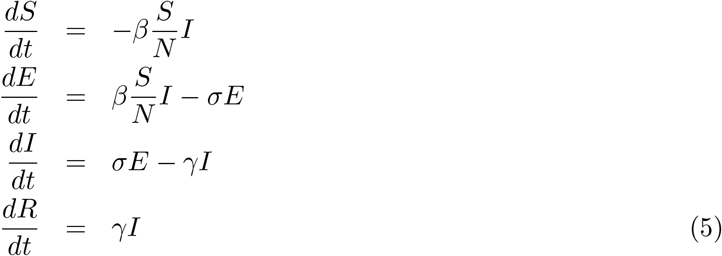

Note that if we combine the second and third equation above we get:

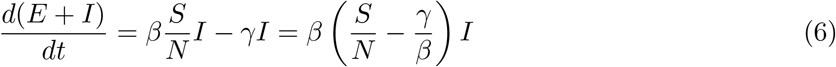

From the above equation we can see that population in the **E** and **I** compartments together can grow with time only when the fraction of the susceptible population is greater than the inverse of the reproduction number:

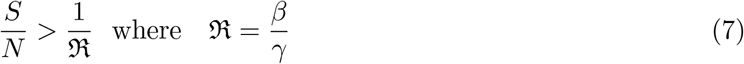

There are many forms of SEIR equations which are in common use (see [BC18, ea20p, ea20u, ea20b, P.20, ea20n]) however, equation (5) is the simplest one and does not include natural deaths. One of the common practices with the SEIR model has been to consider the incubation period 1*/σ* a constant, and estimates it from some other observations. The SEIR modal is quite complex as compared to the SIR model and we cannot find the number of exposed people exactly at time *t* = 0 for evolving the equations and so the approach used to define R no longer works. Thanks to the new generation matrix models [BC18] it is still possible to write R in a close form for this case also.

### 2.3 SIRD model

One of the serious drawbacks of the SIR model is that people who recover and who die are treated in the same way - there are no separate compartments for the dead and recovered people. This drawback can be addressed by separating the compartments for the dead and recovered population as is done in the SIRD model described with the following set of equations (for a detail discussion see [ea20a, Vil20]).

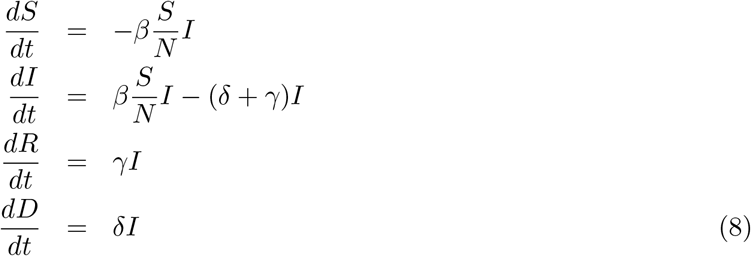

Here a new transmission coefficient *δ* has been introduced which we can identify with the death rate. One of the advantages of the SIRD model is that it has three transmission coefficients *β, γ* and *δ* and we have the data for three time series *I*(*t*)*, R*(*t*) and *D*(*t*) available so it is possible to compute the time dependency of all the three coefficients as well as the reproduction number 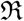.

The aim of any mitigation measures may be one or more of the followings:

1. Lower the contact or infection rate *β*.
2. Lower the mortality rate *δ*.
3. Increase the recovery rate *γ*.

The SIRD model provides us a framework to estimate or fit all these parameters. In one of the coming sections we will discuss how we can reconstruct the transmission coefficients *β, δ* and *γ* as well as 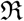 from the data by a direct reconstruction approach.

The basic reproduction rate for the SIRD model can be written in the following way [ea20a]:

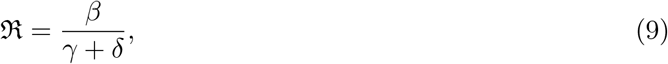

or,

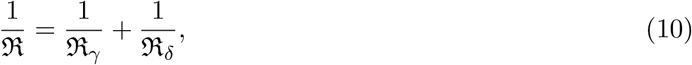

where 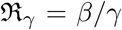 and 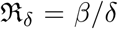. If apart from death and recovery there is some other channel that can lower the population in the **I** compartment, for example if infected people move out from that region with transmission coefficient *η* then we can write:

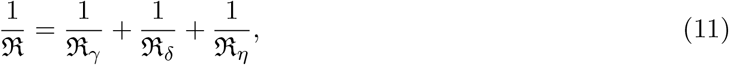

with 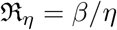. A more realistic model will have multiple compartments (nodes), either connected in series or some branching out from others, with data to constrain the transmission coefficients (edges). Apart from this, realistic models may also require to consider different transmission coefficients of different subgroups (based on age etc.,). Incorporating, all these considerations will lead to very complex models having very less connect with the actual data we have.

### 2.4 Time dependent *β* models

As a pandemic triggers various containment measures [ea20s, DG20, ea20k, SNC20, RR20, ea20c] such as lockdown, social distancing, improved hygiene practices etc., are taken and that lead to transmission coefficients such as *β* becoming time dependent [GPD20, ea20m, FP20, ea20e, ea20i]. Apart from this, the drop in the susceptible population also decreases *β* (see [ea03a, Cob20]).

Lockdown has been one of the most common mitigation measures followed all over the world and, in its extreme form, we can assume that once it starts the contact rate between susceptible and infectious people drops to zero. In general, the lockdown starts on a fixed day *t_l_* and has a duration (time scale) we call *τ* (we will be using both *τ* and corresponding decay rate *µ* = 1*/τ* in the discussion). We can incorporate these two parameters into the modeling of *β*(*t*) in many different ways and a set of three common choices is given below:

1. **Polynomial Suppression** [ea20m]:

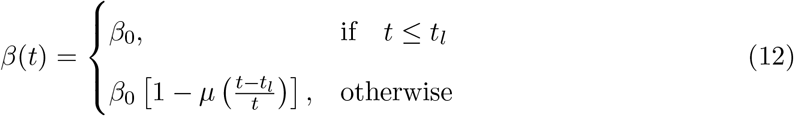

This model is discussed here just for an example and we do not expect the variation of *β*(*t*) as slow as linear one. This expression shows that *β*(*t*) starts with an initial value *β*_0_ and after time *t_l_* it starts decreasing linearly with a constant rate of *µ* = 1*/τ* and finally becomes *β*_0_(1 *− µ*) at *t* = *∞*.
2. tanh **Suppression:**

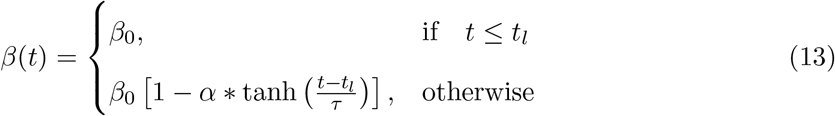 This form of suppression of *β*(*t*) starts with a constant value *β*_0_ at some *t* = *t_l_* and keeps decaying for period represented by *τ* and finally settles to a final value *β*_0_(1 *−α*) as is shown in Figure (3). This can be written in the following way also:

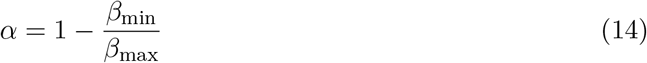 From equation (13) we can also write:

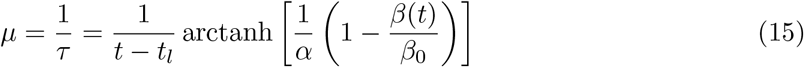 Equations (14) and equation (15) are important to find the priors for *α* and *µ* once we know the priors for *β* and this will be discussed again in §4 and will be used in parameter estimation in §5.
3. **Exponential Suppression** [FP20, ea20o, ea20n]:

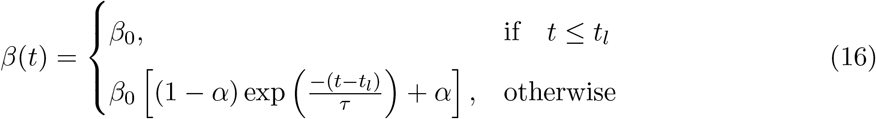 This model is similar to the tanh model and in this case also *β*(*t*) starts from some initial value *β*_0_ and after decreasing for a period and finally approaches to a constant value *β*_0_*α* at *t* = *∞* as is shown in Figure (4). Note that the transmission coefficient *β* may decay with time without any intervention also as is discussed in [ea97] for plants. In this case also we can write:

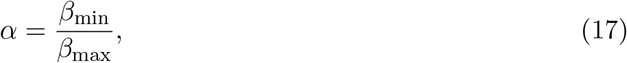

and

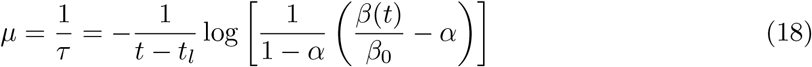 These equations also will be used to find the priors for parameter estimation. In one of the studies [ea20o] it has been argued that even the time of recovery 1*/γ* may also vary with time due to the improvement in medical understanding of the epidemic and facilities and that also can be modeled as an exponential function. There have been other physically well motivated exponentially decaying forms also such as given in [Vil20] in which *β* starts from starting value *β*_0_ and decay with rate 1*/τ* finally becomes *β*_1_.

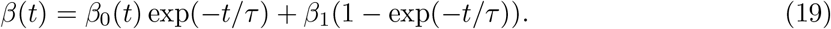 The author argues that *β*_1_ depends on the policy decisions leading to behavioral changes. This model is different from the model we are considering only in the respect that it considers the “lockdown” from the beginning i.e., *t* = 0.

**Figure 3:**
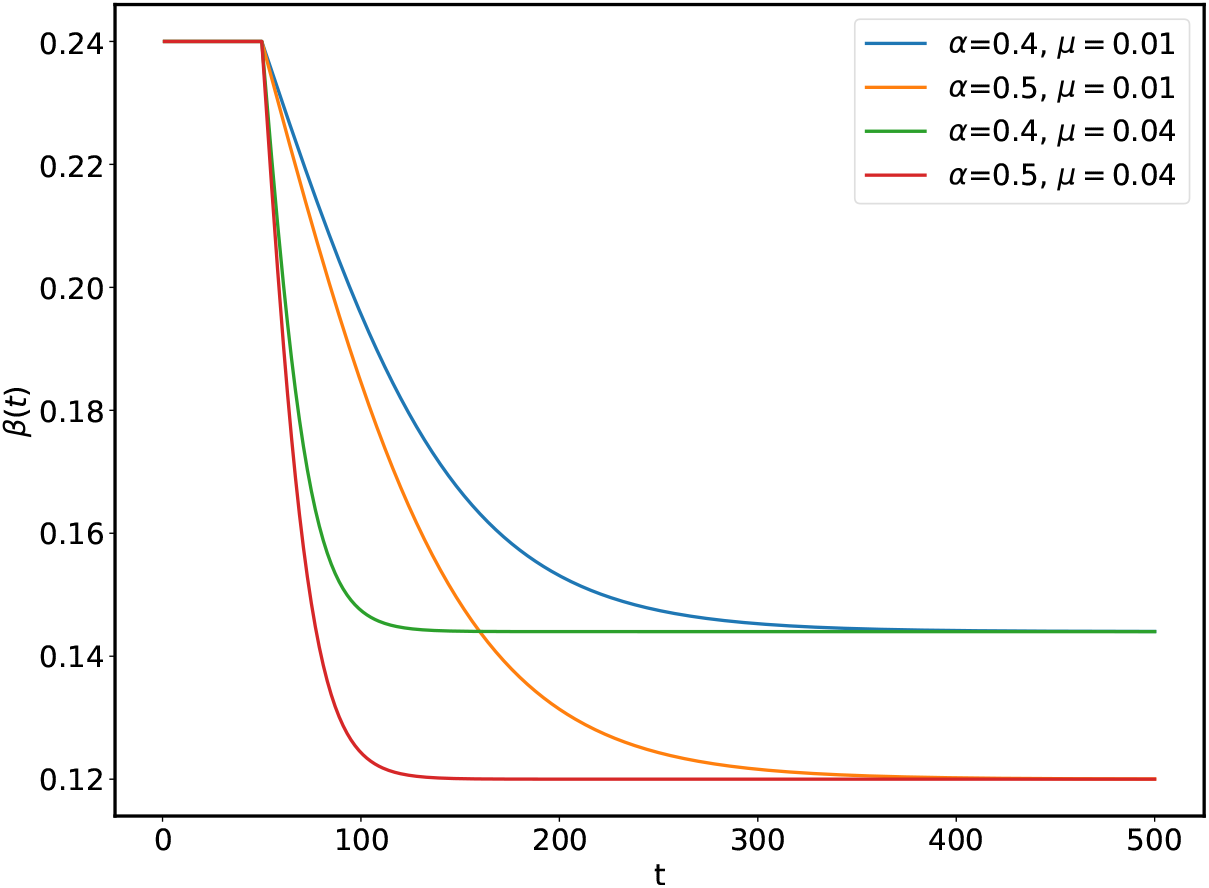
This figure shows the effect of *α* and *µ* on *β*(*t*) for the **tanh** model.

**Figure 4:**
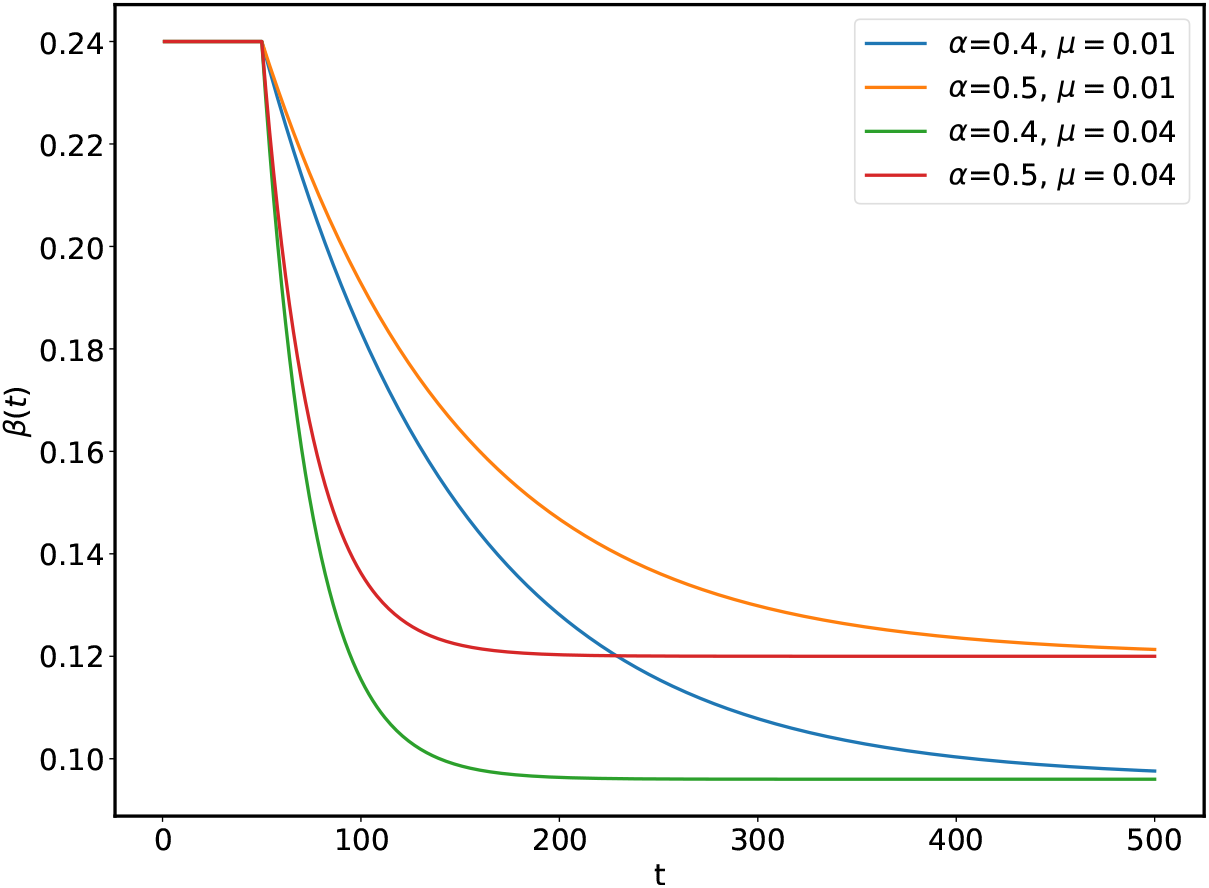
This figure shows the effect of *α* and *µ* on *β*(*t*) for the **exp** model.

The time dependent *β* models as are discussed above and shown in Figure (5) share a common property that before a certain time *t_l_*, that we can identify with the day on which lockdown starts, *β* has a constant value *β*_0_ and after that it starts decreasing with a rate that depend on the parameter *µ* = 1*/τ*. The effect of the suppression in *β* is controlled by the parameter *µ* and for its zeros values all the models become constant *β* models. From Figure (5) we can conclude that different models can lead to the same amounts of “flattening” of the curve with a different choice of parameters so there is no preferred model for the suppression.

**Figure 5:**
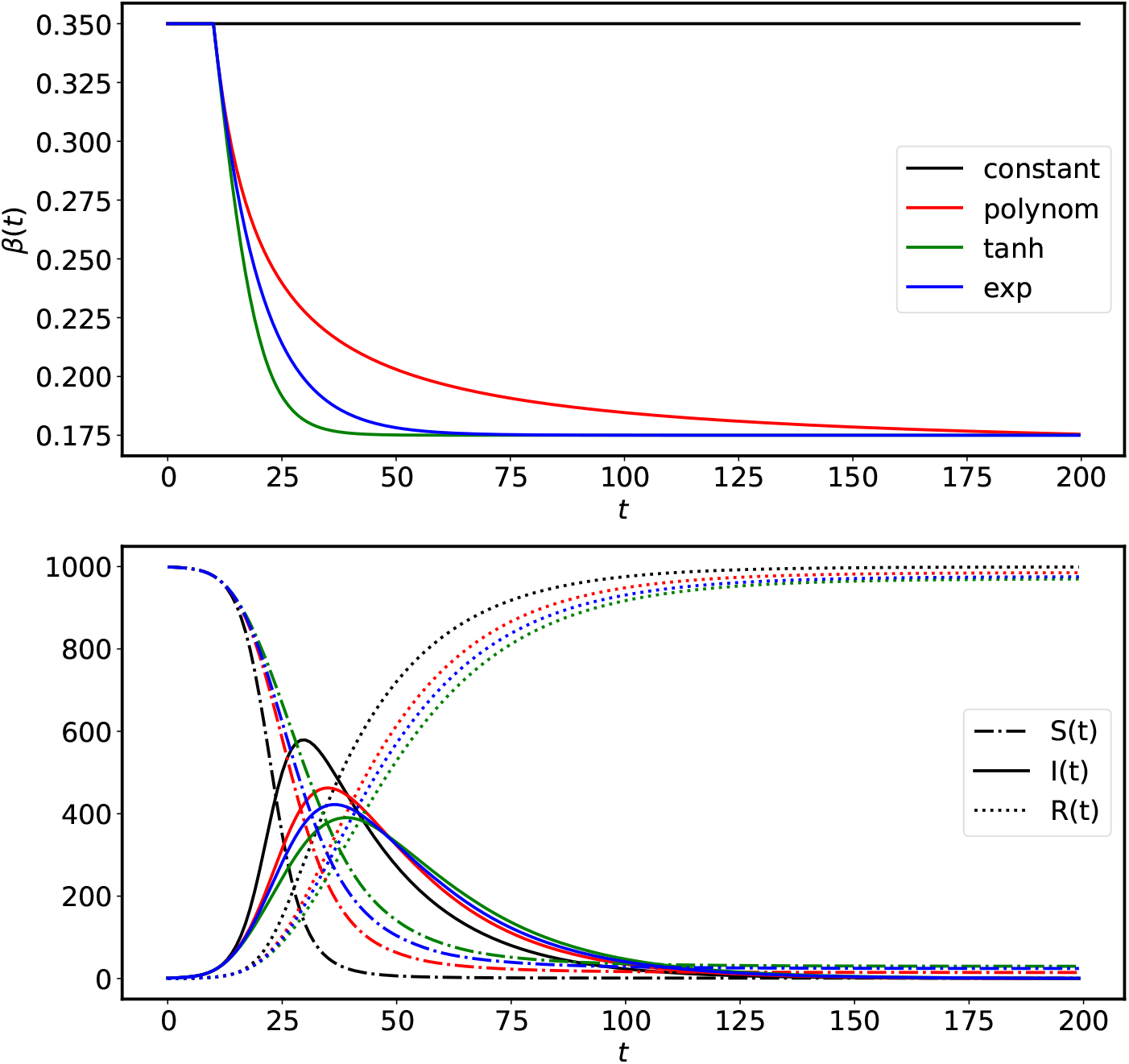
The top panel in the figure shows the *β* suppression models as are discussed in the text. The solid and dashed line in the bottom panel show the number of infected people *I*(*t*) and the number of recovered people *R*(*t*) for the SIR model corresponding to different *β* models. From the bottom panel we can see that all the models have similar effects, however, for the tanh case the suppression in the peak of *I*(*t*) is maximum. This figure is for an illustration the values of parameters have been chosen carefully to highlight the effects.

The SIR model with constant transmission coefficients is applicable only in the situation when the pandemic is let to grow without any intervention. In the real world once a pandemic starts interventions of different kinds (social, medical etc.,) are considered to reduce the rate at which the the epidemic spreads. These interventions can be easily taken into account by considering a time dependent (decaying) growth rate (*β*). As we can see from the above figure that a decaying (exponentially) *β* helps to contain the disease by lowering the height of the peak as shown in Figure (5).

## 3 Data

The primary data for Covid-19 is in terms of three times series for the count of confirmed 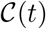, recovered 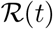 and dead 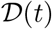, persons for every country. By definition all the three times series are non-decreasing functions of time, as are shown in Figure (6) for Italy. The data for Covid-19 is provided by the John Hopkins University [ea20g, Gar20a, Gar20b] and is updated on a daily basis and interactive tools are also provided for data exploration. The worldometer website [Wor20] and **our world in data** [iD20] also provide an up to-date data for most countries with some extra information, such as the numbers for the active cases, critical cases and the test conducted for a million population of the country.

**Figure 6:**
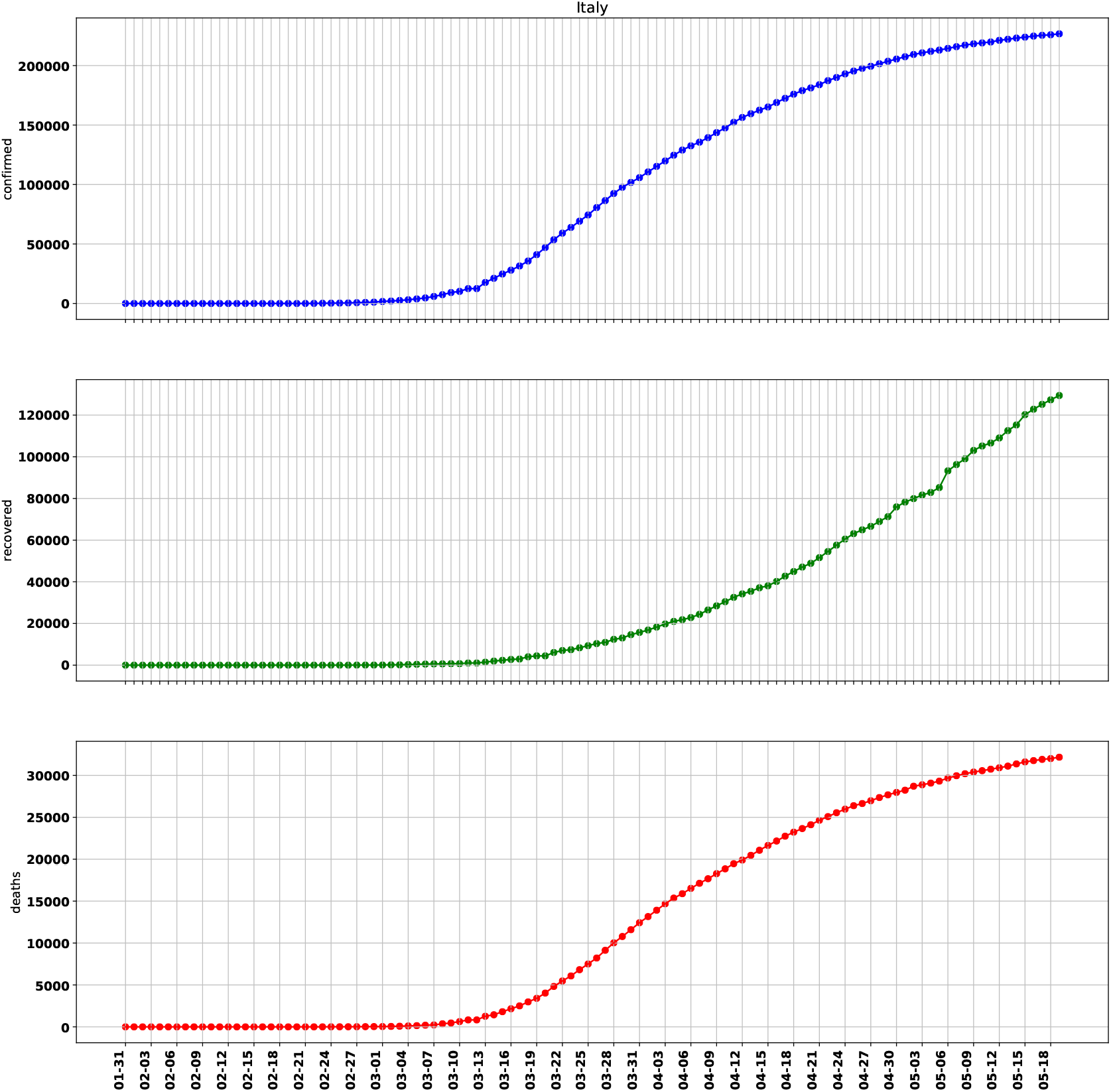
The total number of confirmed, recovered and dead case for Italy.

The time series which are shown in Figure (6) grow exponentially in the beginning and then settle to a slower power law growth. If we look at the daily new cases, as shown in Figure (7), they look like broad peaked functions with increasing fluctuations around the the current value. Time series for a set of countries used in the analysis are shown in the Figure (8).

**Figure 7:**
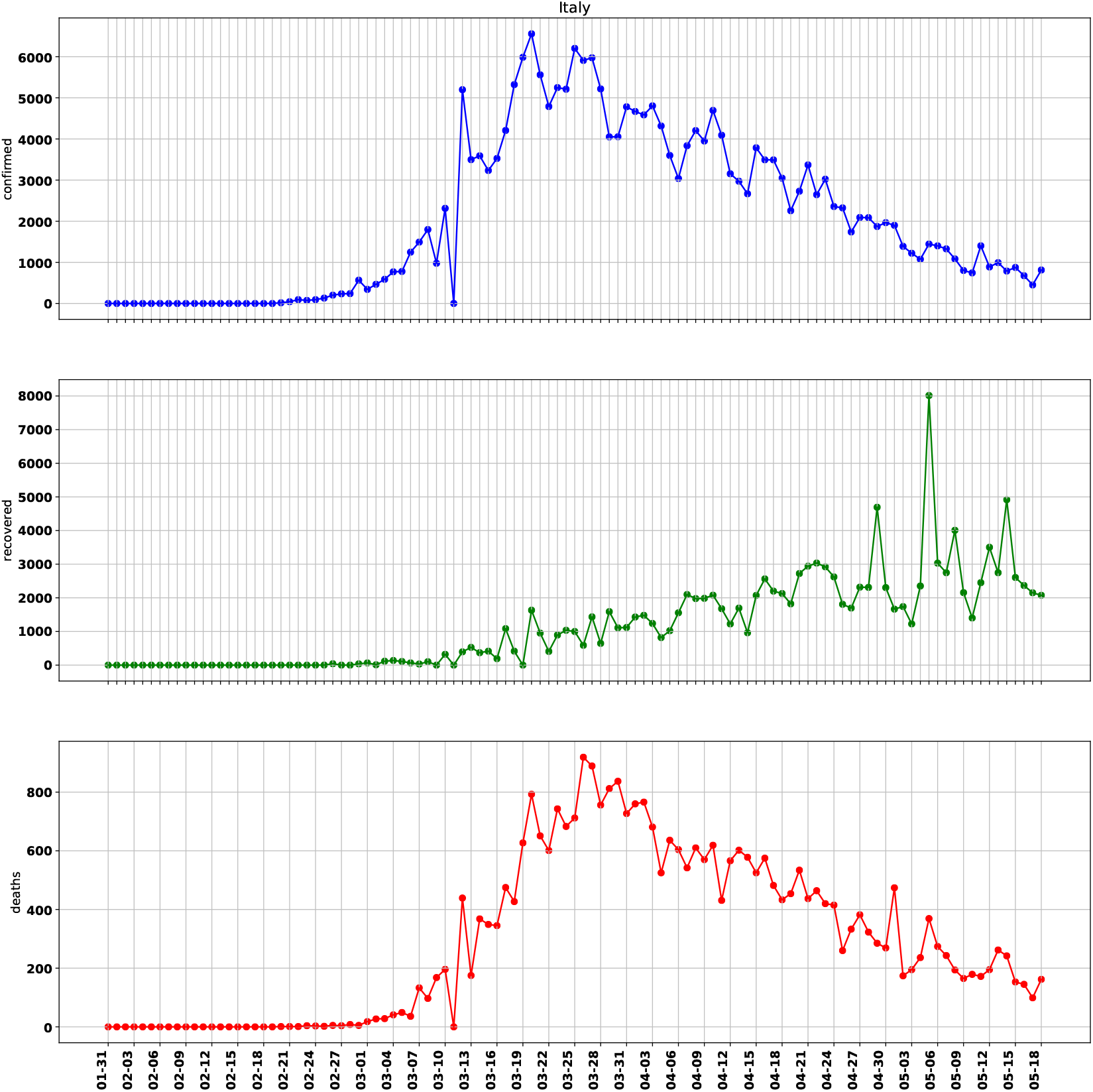
The number of new confirmed, recovered and dead case for Italy.

**Figure 8:**
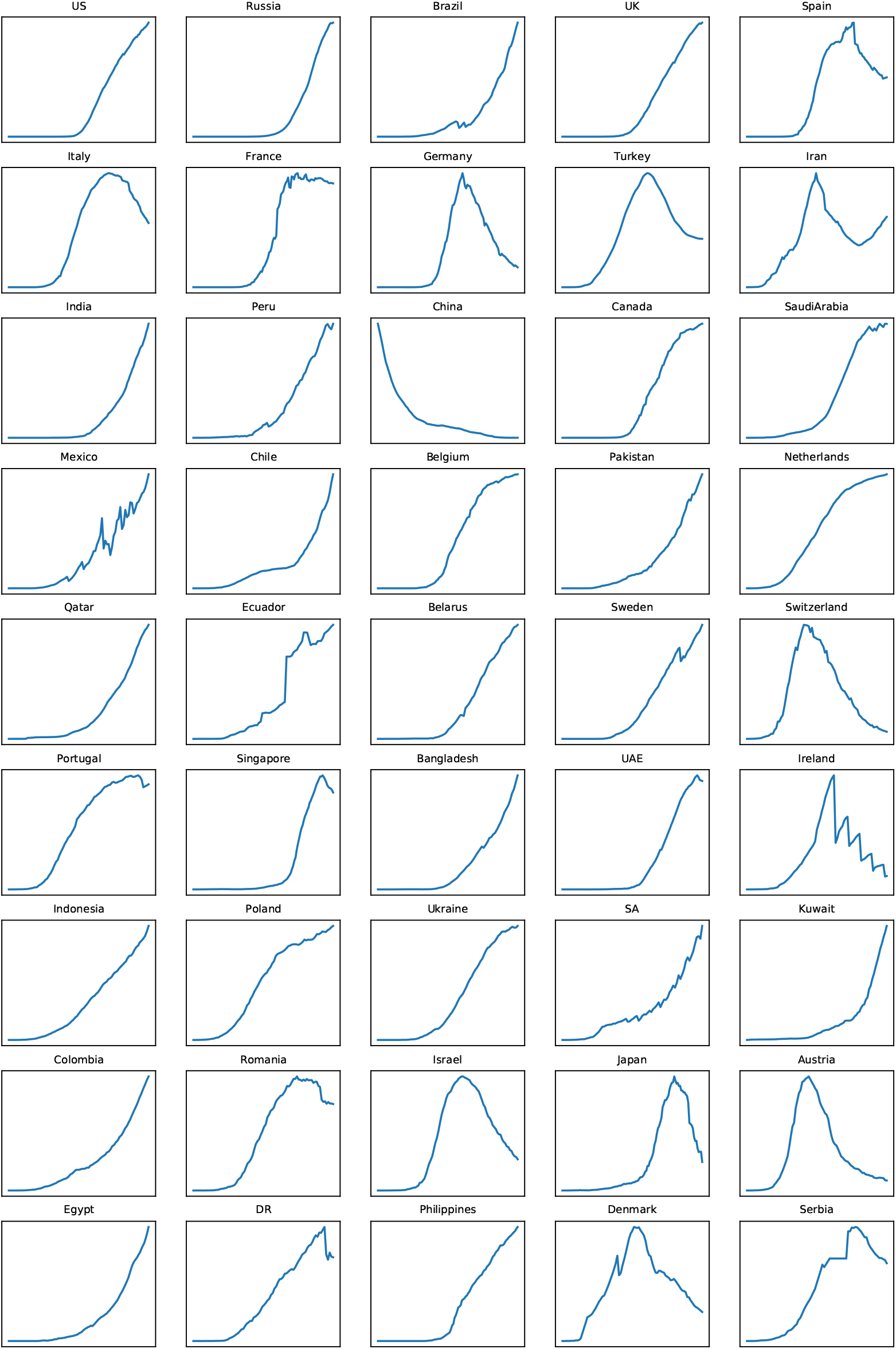
Time series for a set of countries we used for modeling.

There are many factors, known and unknown, which determine the behavior of these time series, such as the nature of the diseases/virus, the health profile of the population, availability of medical infrastructure, age-distribution, social mixing, personal hygiene and may be geographical location and genetic makeup of the population etc. Compartmental models, such as SIR, SIRD, SEIR etc., help to simplify the problem by replacing a large number of controlling parameters with a very small set of well motivated parameters - the transmission coefficients.

The compartmental models predict that how the population in different compartments change with time. In the SIR model the population in **S** and **R** compartments can only decrease and increase respectively, however, in the **I** compartment it can increases as well as decreases. On the onset of the epidemic almost everyone is in the **S** compartment with a very small fraction in **I** and no one in the **R** compartment. At the end of the pandemic everyone is in the **R** compartment with no one in **S** or **I** compartment (see Figure (1). The same happens for SIRD model also where the decrease in **I** compartment happens due to recovery (R) and deaths (D).

The time series *I*(*t*) for the population in compartment **I** can be obtained by subtracting 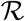 and 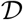 from 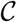.

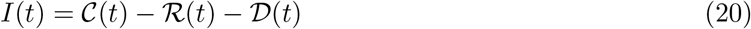

For a set of countries the time series of *I*(*t*) are shown in Figure (8). The time series 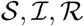 and 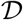 have very weak dependency on each other - the only constrains they have to satisfy is that the sum total of the population in different compartment must add to the total population. The number of people *I*(*t*) in the **I** compartment at time *t* does depend on all the three transmission rates *β, γ*, and *δ*, therefore it is a good measure which we can fit to a compartmental model, such as SIR or SIRD and can get constraints.

**Table 2:**
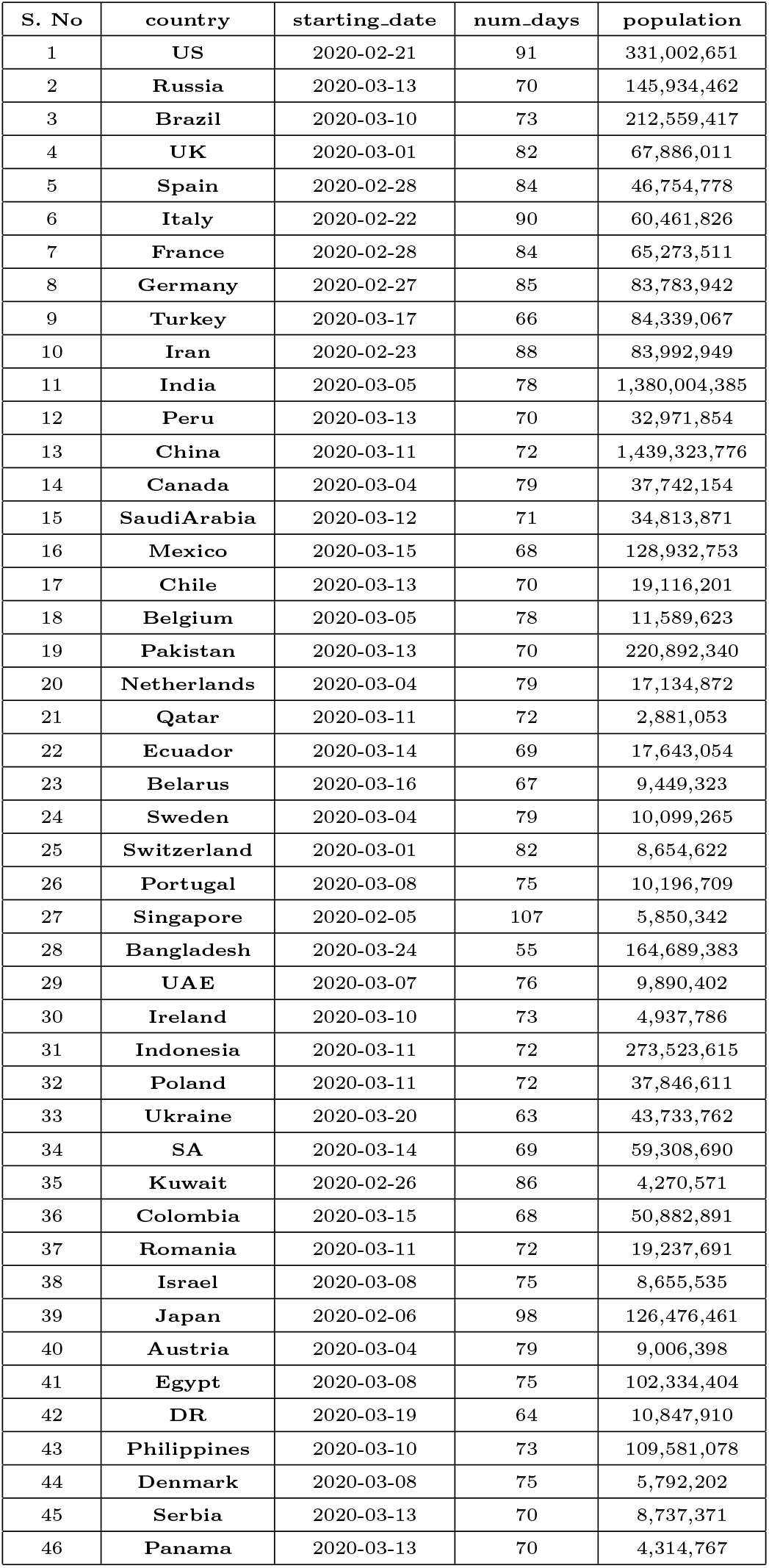
A summary for the countries used for modelling

## 4 Reconstruction

In this and next section we present the main results of the study in the form of demonstrating a reconstruction procedure for the time dependent transmission coefficients *β*(*t*)*, γ*(*t*)*, δ*(*t*) and the effective reproduction number 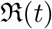. We consider an example of Italy for this procedure. Note that this approach is common and can be used to understand the variation of the transmission coefficients with time as a result of interventions. The main advantages of this approach is that there are no parameters to adjust and so the results are easy to reproduce.

The approach we use here is similar to as used in [ea20e, GPD20]. In this approach the evolution equations are written in a discretized form as shown in equation (21).

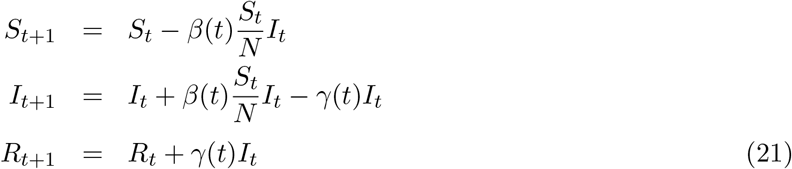

From the third equation we can write:

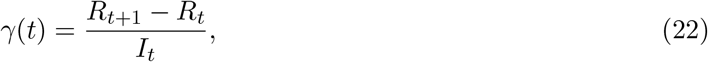

and using this and second equation from (21) we get,

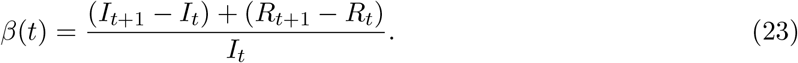

Note that by definition *R_t_*_+1_ *≥ R_t_*, so *γ*(*t*) *≥* 0, however, we may have *I_t_*_+1_ *≤ I_t_* also, *β*(*t*) may become negative also once the population in the compartment **I** starts decreasing.

Here an important assumption is being made and that is the fraction of susceptible population *S/N* is close to unity which may be true at the beginning of the epidemic. Once we have expressions for the time dependent *β* and *γ* we can also written an expression for the time dependent reproduction number in the following way:

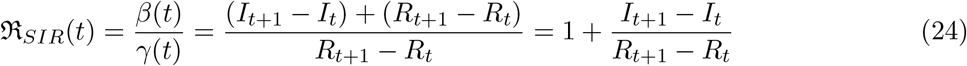

Following the similar procedure we can write the SIRD equations in the following discretized form:

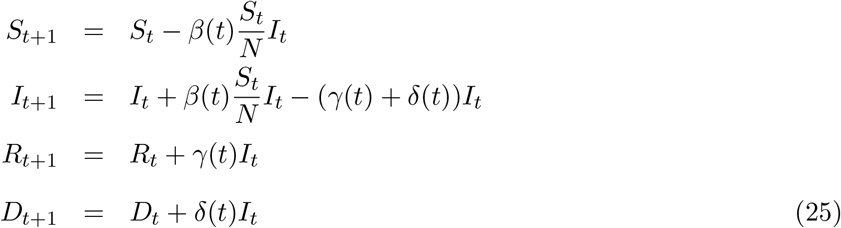

From these questions we can write:

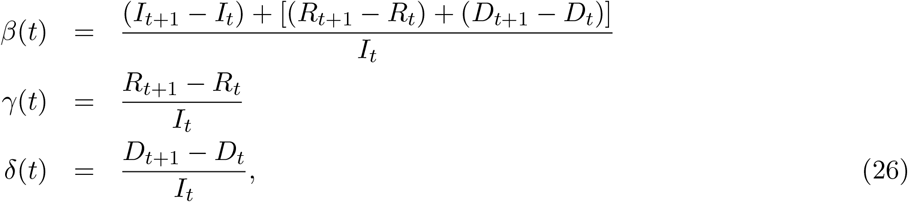

and can write the expression for the reproduction number:

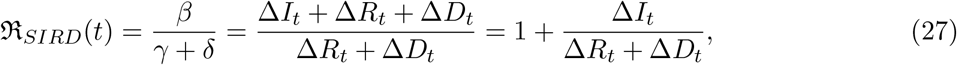

where ∆*X_t_* = *X_t_*_+1_ *− X_t_* with *X* = *I, R* and *D*. This equation is identical to equation (24) if we do not count dead and recovered separately i.e., replace ∆*R_t_* + ∆*D_t_* with ∆*R_t_*. One of the interpretations of *R* is that it is a ratio of two rates and so in case we are interested finding out two separates measures for *γ* and *δ*, we can also write:

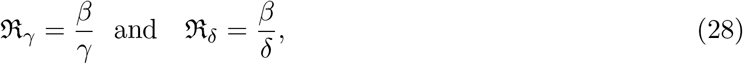

and so,

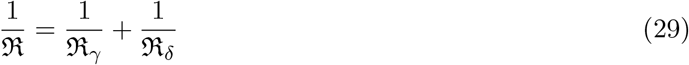

The procedure as discussed above can be used to know the variation of the transmission coefficients *β, γ, δ* and effective reproduction 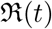 with time. In order to follow this procedure we need to abandon first few data points which have very high noise. As explained above occasionally we may also have negative values of R(*t*).

In figure (9) we show the reconstruction for *β*(*t*)*, γ*(*t*) and *δ*(*t*) for Italy with SIRD. From this figure it is clear that all the three transmission coefficients vary with time and the variation of *β*(*t*) is maximum. The same trend is observed for other countries also. In the next section will fit a parameterized form of *β*(*t*) which we obtain from the model-fitting to the data shown in the figure.

**Figure 9:**
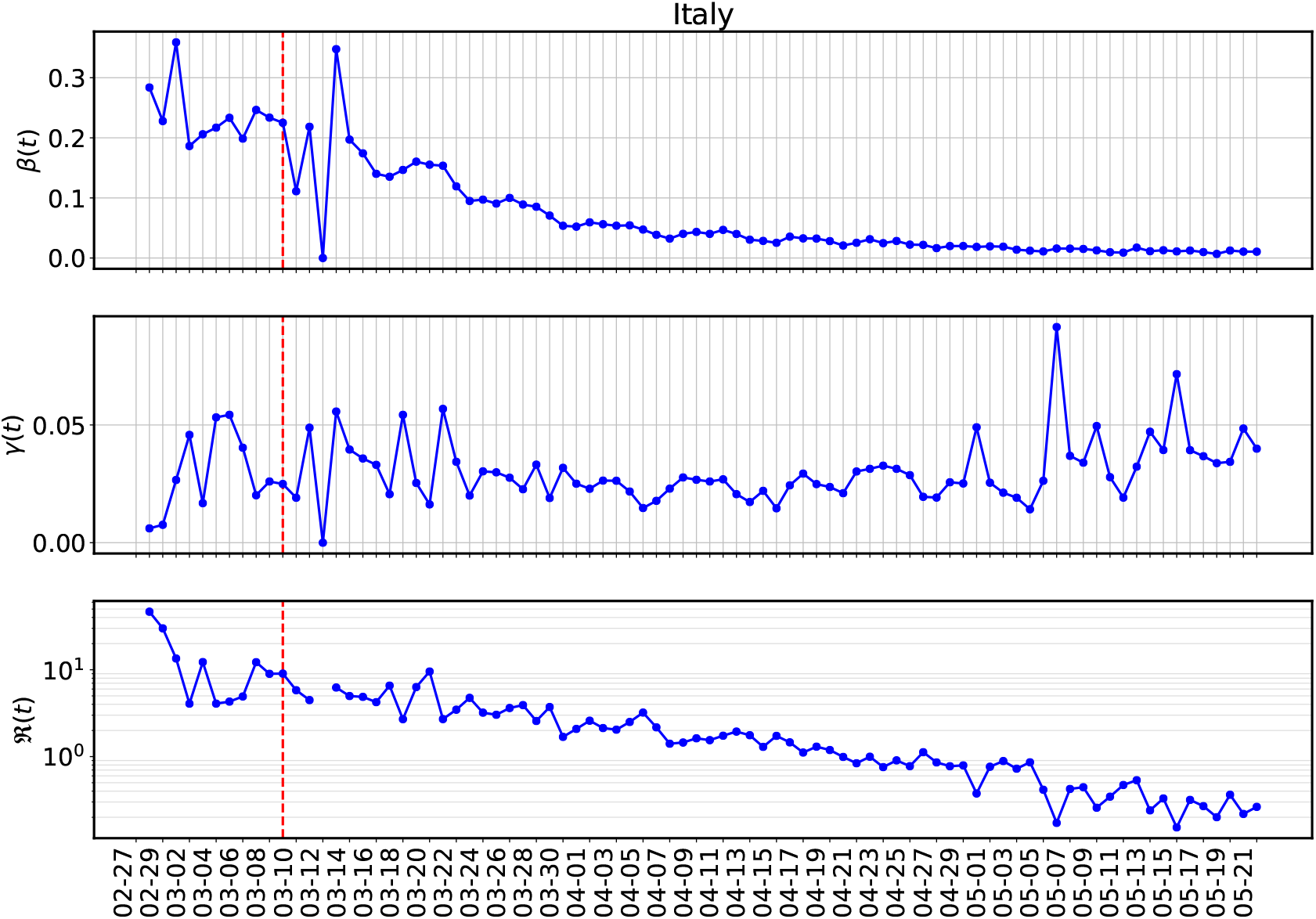
Reconstruction of the transmission coefficients *β*(*t*)*, γ*(*t*) and *δ*(*t*) with the SIRD model for Italy.

**Figure 10:**
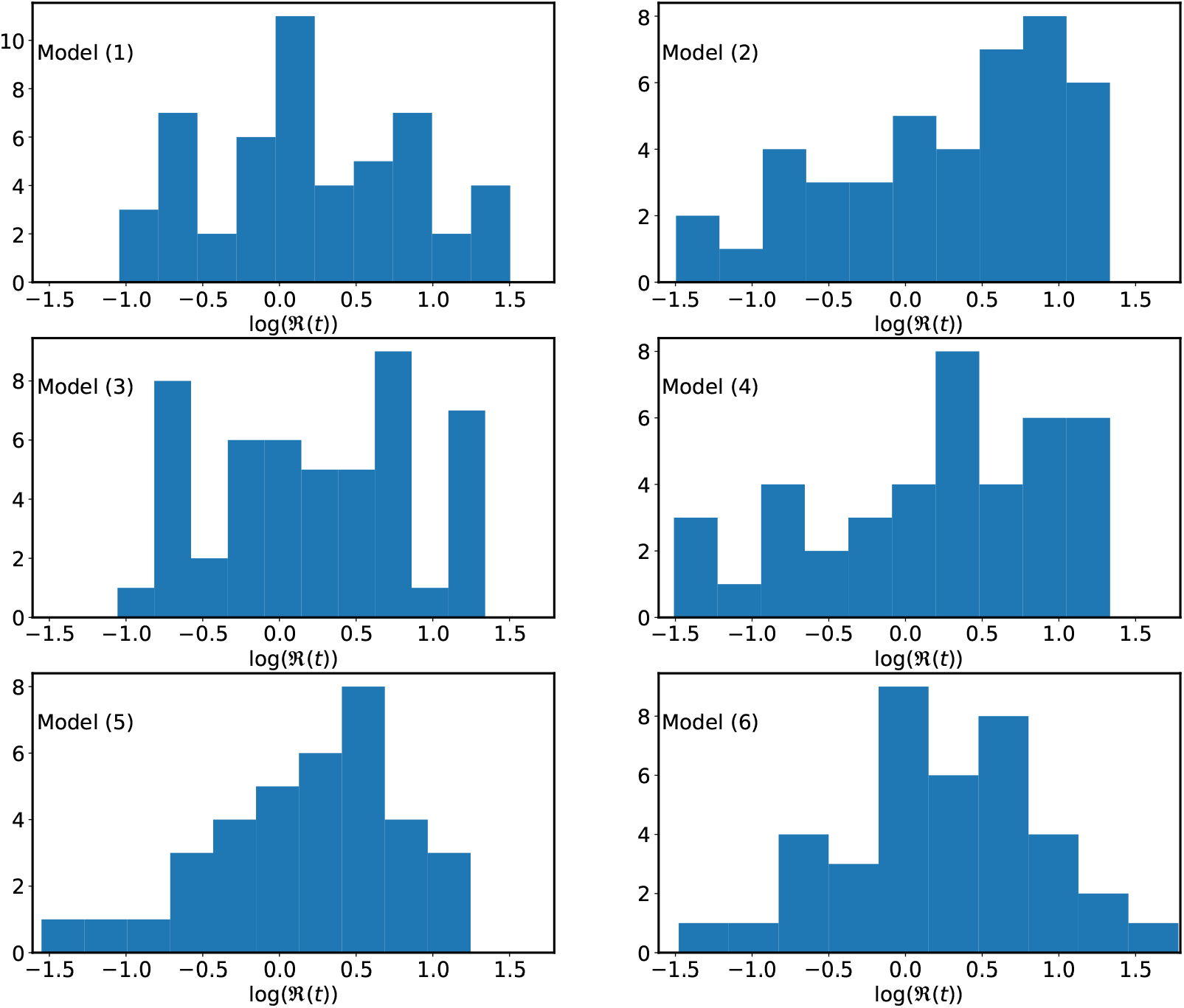
Histogram of the effective reproduction number 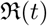 with different models.

Once we have reconstructed *β*(*t*)*, γ*(*t*) and *δ*(*t*) we can easily get 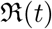 for SIR and SIRD model.

### 4.1 Priors

One of the import uses of the reconstruction procedure we have discussed here is to find the priors (minimum, maximum and best fit) values for the parameters to be fitted. Once we have the estimates for *β*(*t*)*, γ*(*t*) and *δ*(*t*) from the above procedure we can easily find *X*_min_*, X*_max_*, X*_0_, values (with *X* = *β, γ, δ*. Here, *X*_0_ is the approximate point for the parameter that is needed in many optimization procedure which iteratively find the solution. Since in the present work we use parametric form of *β*(*t*), so we need priors for the parameters of *β*(*t*) i.e., *β*_1_*, α, µ* and *τ* which can find from the reconstructed *β*(*t*) (see §2.4 for detail).

## 5 Model fitting and parameter estimation

We consider a set of six compartmental models, three belonging to the SIR and three to the SIRD class. The models are different from each other in terms of the choice for the epidemiological class (SIR or SIRD) or the model for the contact rate *β*(*t*) (see §2.4 for detail). A summary of the models is given in Table (3).

**Table 3:** A summary of the models being considered for the analysis

| Model | Epidemiological model | $\beta(t)$ Model | Parameters |
| --- | --- | --- | --- |
| 1 | SIR | exp | $(\beta, \gamma), (\beta_0, \alpha, \mu, t_l)$ |
| 2 | SIR | tanh | $(\beta, \gamma), (\beta_0, \alpha, \mu, t_l)$ |
| 3 | SIRD | exp | $(\beta, \gamma, \delta), (\beta_0, \alpha, \mu, t_l)$ |
| 4 | SIRD | tanh | $(\beta, \gamma, \delta), (\beta_0, \alpha, \mu, t_l)$ |
| 5 | SIR | exp | $(\beta, \gamma), (\beta_0, \mu)$ |
| 6 | SIRD | exp | $(\beta, \gamma, \delta), (\beta_0, \mu)$ |

Note that in the model (5) and (6), *β*(*t*) starts decaying from the very beginning (in place of starting from a particular day representing the date of the lockdown) with a constant rate *µ*.

In any fitting procedure the choice of the loss function depends on what we wish to fit. In the common least square fitting we use the sum of the squares of the offsets as the loss function. However, there is a problem here with the data we have for that choice. The time series we wish to fit have small values at the beginning and very large values at the later stage, so the the fitting is biased towards the points which have large values. One of the solutions for this could be to fit the log of the time series but then the fitting becomes biased toward small values, in the beginning (or later stage when the peaks falls).

We decide to use the loss function of the ordinary least square which fits the data points close to the peak (having higher values) more accurately than other data points. We found this useful for the following two reasons:

1. The peak in the time series is an important feature, in particular its location and height, therefore any loss function biased towards it is justified.
2. For the short term predictions only the data points close to the dates of prediction is important, so using a loss function that fits later points (having higher values) more accurately than the noisy data points in the beginning is favorable.

The loss function which we used for fitting the data to SIR and SIR models is given below.

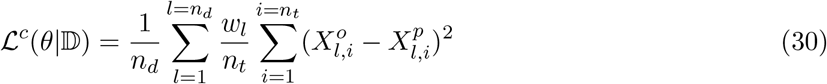

The variables used in the above equations are defined in table (4):

**Table 4:** A summary of the variables

| Variable | Name | Description |
| --- | --- | --- |
| $\mathbb{D}$ | Primary Data | $\mathcal{C}, \mathcal{R}, \mathcal{D}$ |
| $\theta$ | Fitting parameters | As defined in the last column of Table (3) |
| $X_l^o$ | Fitting data | $I, R + D$ for SIR and $I, R, D$ for SIRD |
| $X_l^p$ | Predicted data | prediction from the model |
| $n_t$ | Days | Number of days for which we have data |
| $n_d$ | Number of time series used | 2 for SIR and 3 for SIRD |
| $c$ | country | label for the country |
| $w_l$ | weight | Used for fitting multiple time series together |

Once we have estimated the fitting parameters of our model we can find a smooth representation of *β*(*t*) using the best-fit values of the parameters *β*_0_*, α, µ* and *t_l_* as is shown in Figure 11 for Italy with SIR model and **exp** model for *β*(*t*). The first few data points are very noise and so are not shown in the figure. The similar exercise is also done for 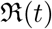 in Figure (12). One of the shortcomings for the case of 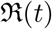 is that we have assumed that *γ* as a constant although the data does show the variation of *γ* and *δ* with time (see Figure (9). We have also shown the beta model fitting for India and US in the Figure (13) and (14).

**Figure 11:**
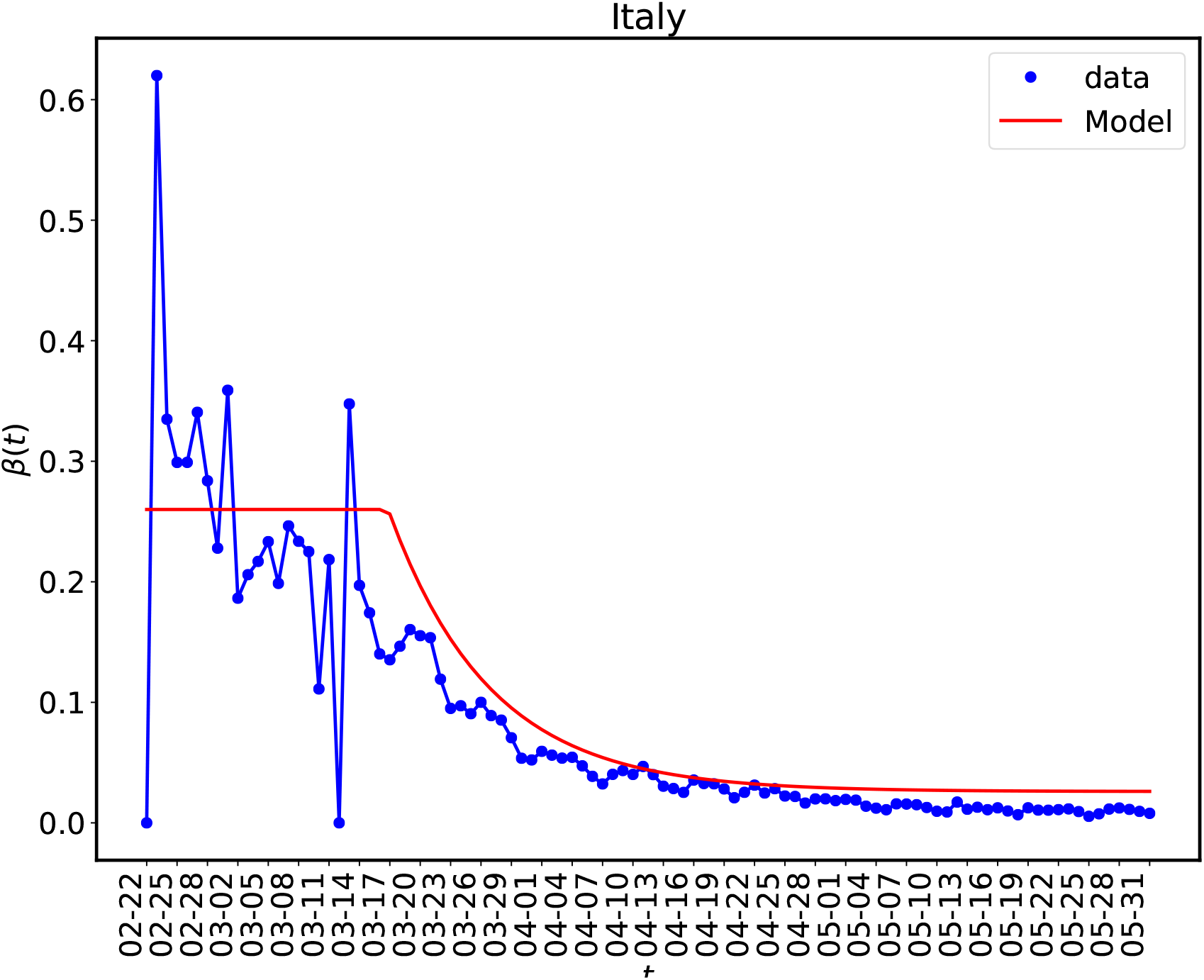
In this figure we have shown the reconstructed *β*(*t*) and a smoother version which we have obtained by model fitting for the SIR case with **exp** model for *β*(*t*). Similar exercise can be done for other countries also. Since the data points are very noise so it does not make much sense to use more complex model for *β*(*t*) that what has been used here.

**Figure 12:**
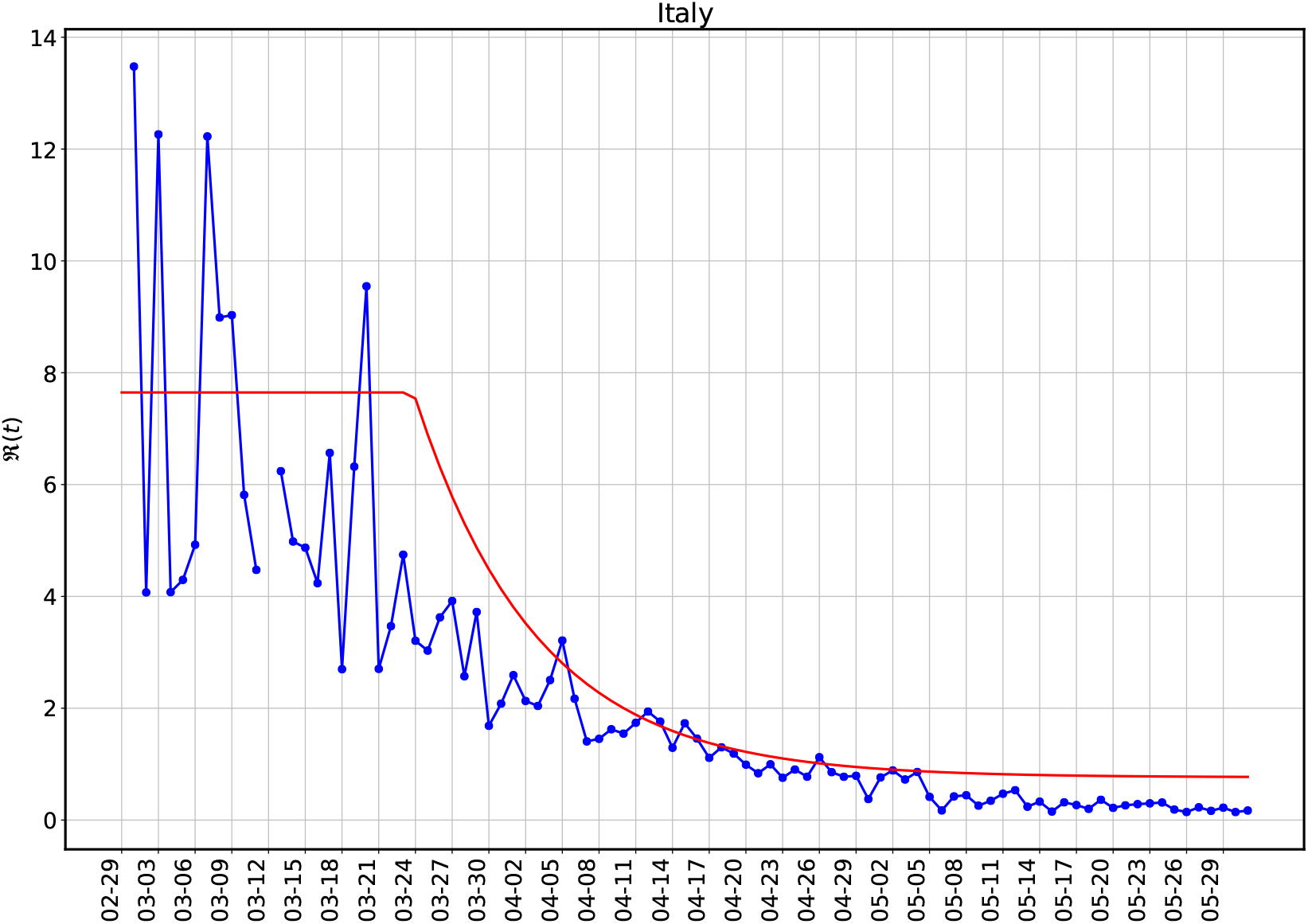
Reconstructucted effective reproduction number 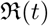for the same set of parameters as are used in Figure (11)

**Figure 13:**
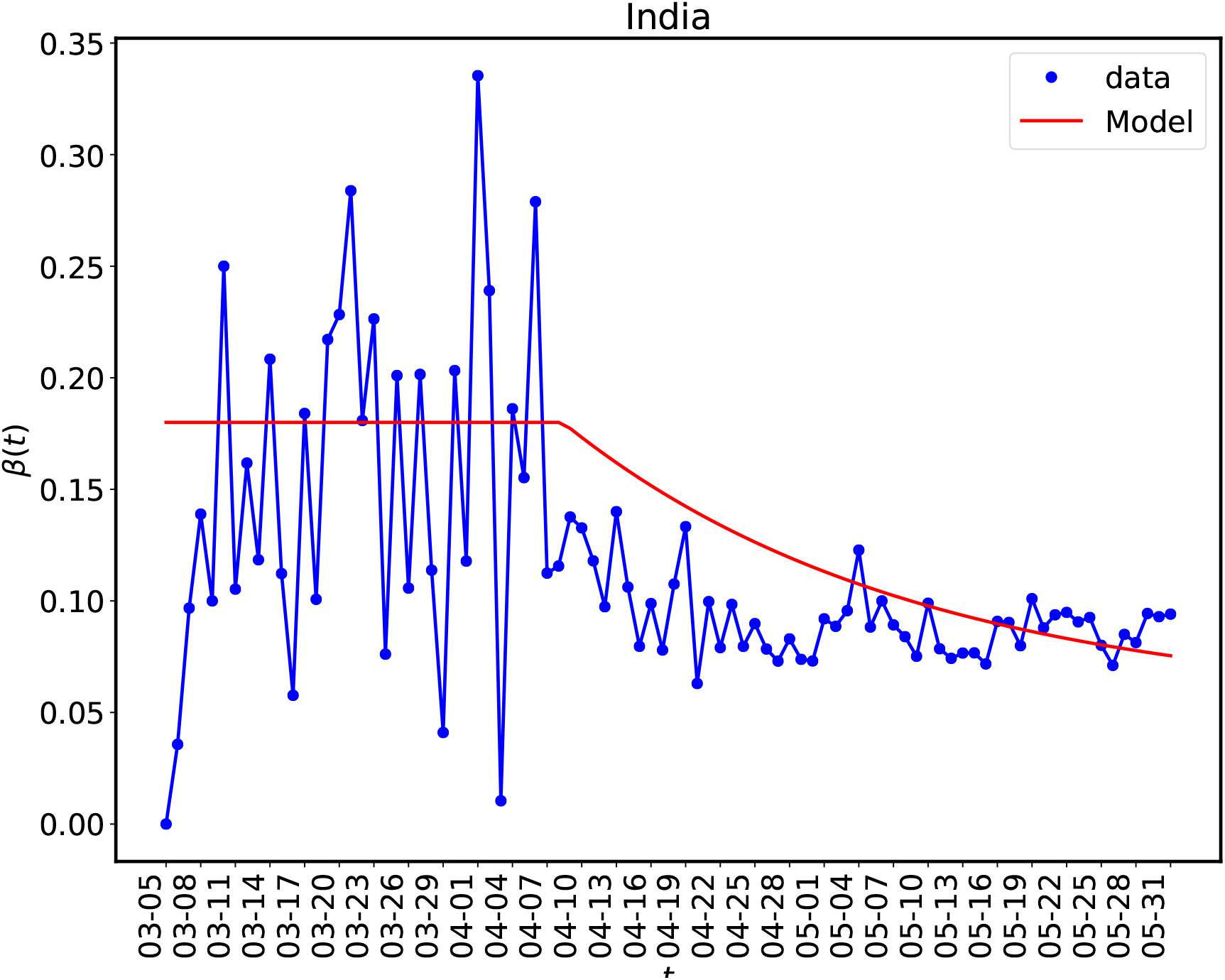
The same as in Figure (11) for India.

**Figure 14:**
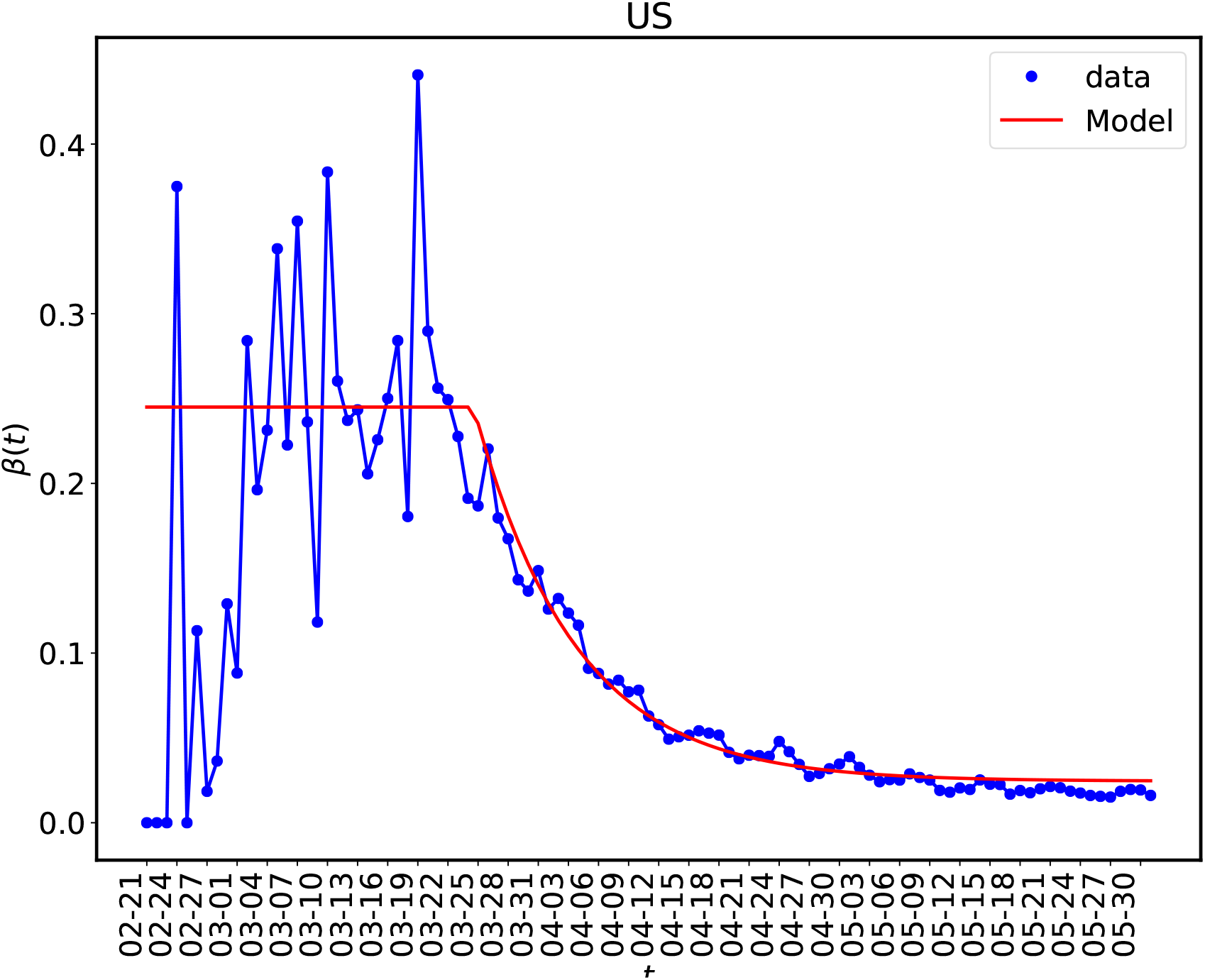
The same as in Figure (11) for US.

For SIR and SIRD models we fit multiple time series together so we must weight the sum of the squares of the offsets for different time series since they have very different values - the value of *I*(*t*) is generally few orders of magnitude higher than *R*(*t*) and *D*(*t*). We use the following weights for this purpose:

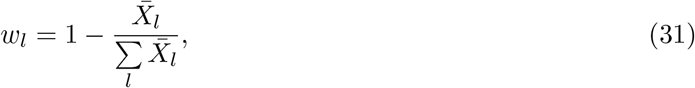

where 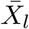 is the average of the time series *X*(*t*).

We use the **solve_ivp** and **minimize** modules from the **Scipy** [Sci20] for integrating the differential equations and minimize the cost function respectively. The loss function given by equation (30) represents the Root Mean Square Deviation (RMSD) and we use its final value as a measure of the goodness of the fit.

A list of fitting parameter for the different models is given in Table (3). For the SIR class of models, model (1) and (2) we have five fitting parameters named, *γ, β*_0_*, α, µ, t_l_* and for the SIRD models, model (3) and (4) we have six fitting parameters named *γ, β*_0_*, α, µ, t_l_* and *δ*. As we can notice that for model (1) to model (4) four of the parameters are associated with *β*(*t*) and for model (5) and (6) the variation of *β*(*t*) is controlled by just two parameter - *β*_0_, the initial value of *β* and its decay rate *µ* = 1*/τ*.

The best fit values of the fitting parameters with their 90 % CI (standard deviation) as well as the median values are given Table (5). The tables also give the estimate for the effective reproduction number 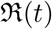 which is a derived quantity here. Note that for computing 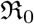 we have extrapolated the value of 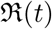 to the last date for which the data is being used here.

**Table 5:**
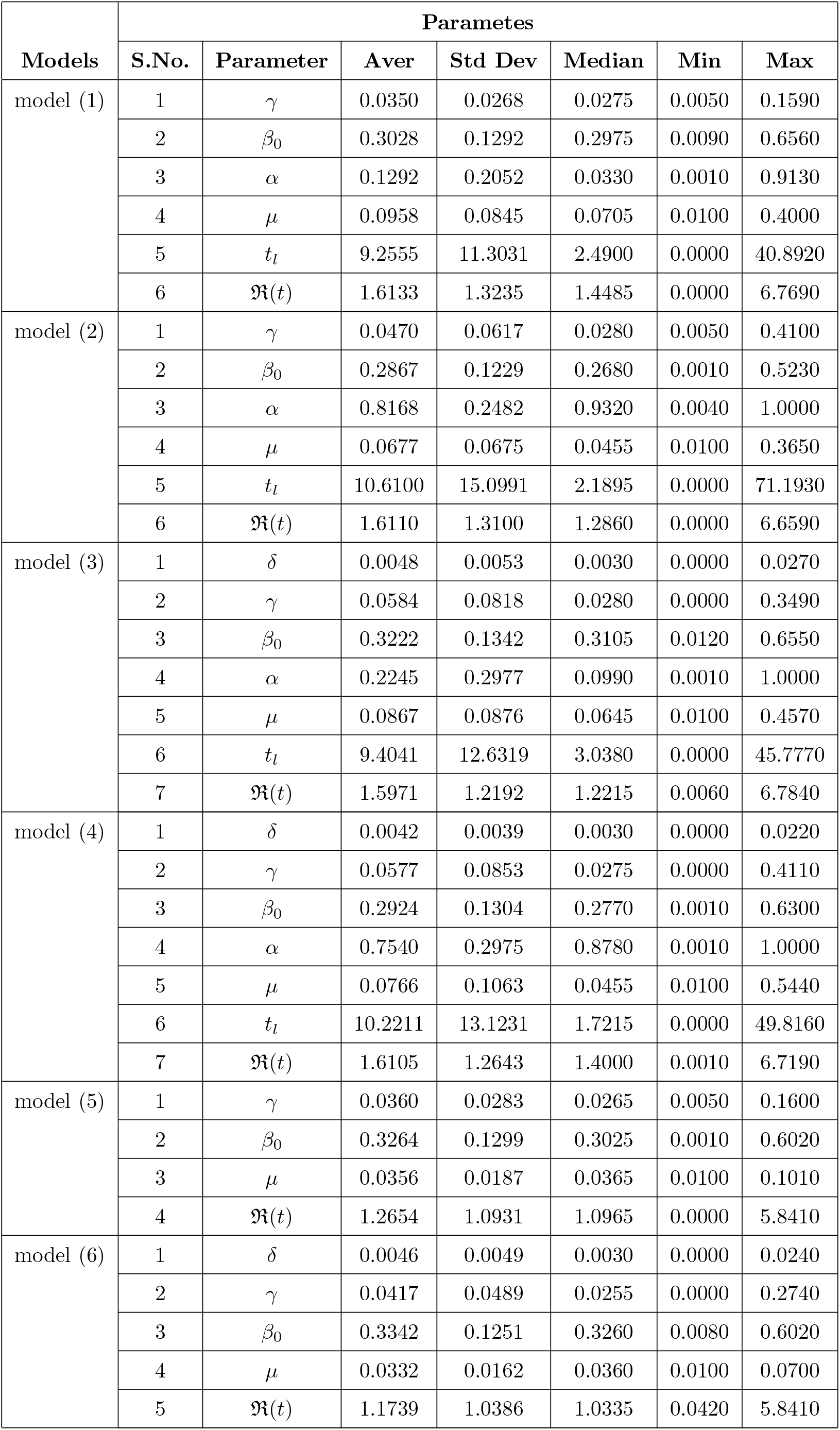
A summary of the fitting parameters for all the models.

A histogram of the effective reproduction number for the different models being considered is shown in Figure (10) and detail values of that for different countries, which include the average values as well as 90% CI (StdDev), are given in Table (6).

**Table 6:**
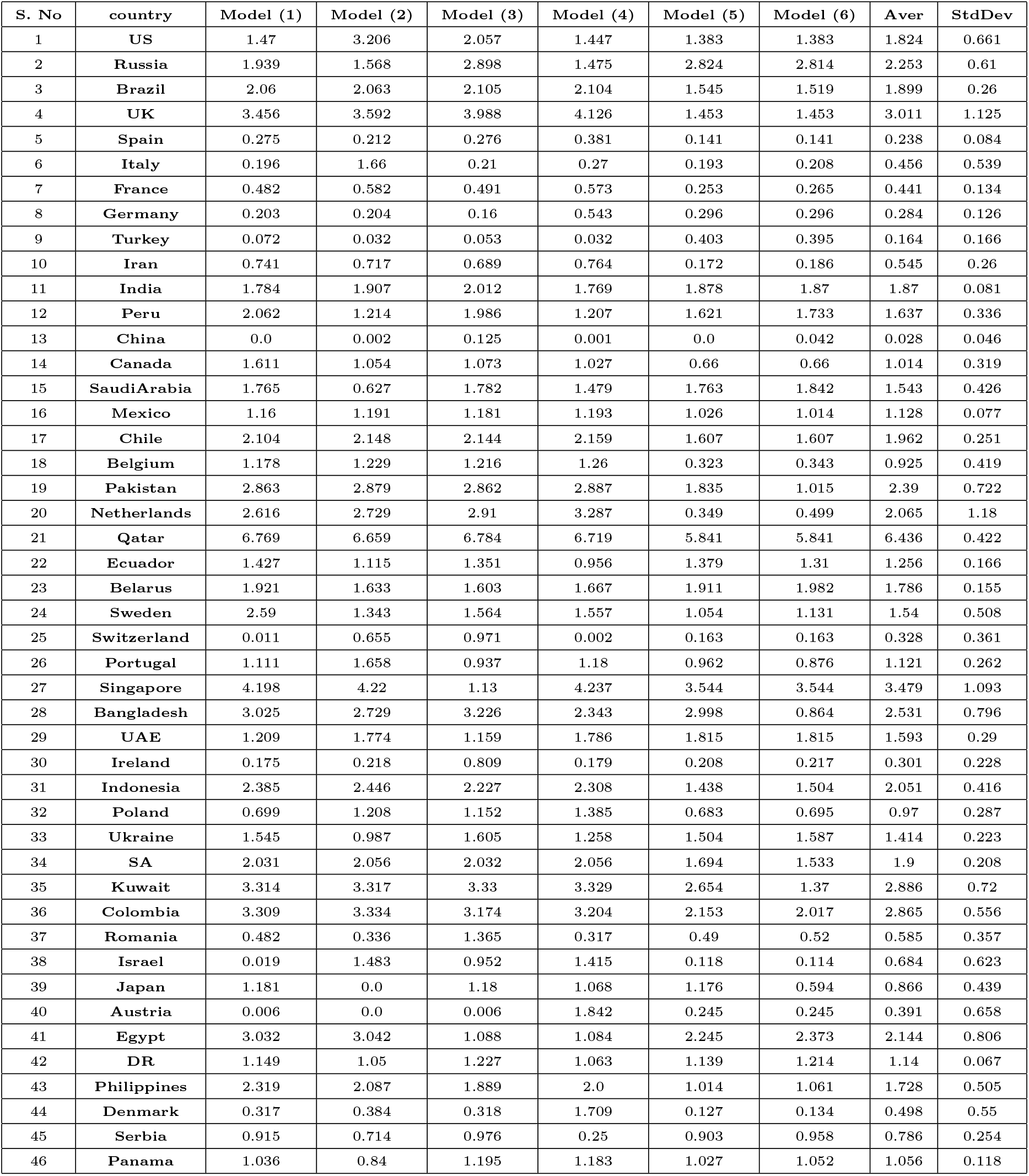
Effective reproduction number 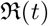 on, May 31, 2020, for countries with different models

## 6 Summary and conclusions

Covid-19 is a global crisis and understanding its impact on different systems of the modern human life (medical, social, economical etc.,) and the responses presented is an important exercise to carry out. We understand that despite being a global phenomenon, the impact of Covid-19 in terms of the loss to life and the resourced being exhausted depend on the local conditions as well as on the mitigation measures taken locally. However, we believe that the global picture of the crisis does help to plan and take policy decisions at the local scale also.

Full understanding of any pandemic, in particular like Covid-19 which does not have any other examples in the history (in terms of the scale and impact), may become available only when it is over and the facts and figure presented here may have very short life. However, we still believe that any quick timely insight may help a lot in terms of the planning for the worse. Knowing very well that all the mathematical models are wrong but some are useful, we believe that mathematical models which are presented in this work may help to develop some insight about the crisis. A brief summary of the work presented here is as follows.

In §1 we have given a very brief introduction of the problem being addressed and reviewed some of the key works about Covid-19 which motivated the present work. A brief introduction of the mathematical framework used in the work in §2, in particular we have review a set of compartmental models SIR, SEIR and SIRD in §2. We have also discussed a set of of parametric models for one of the transmission coefficients *β*(*t*), in §2.4. We have discussed the data being used in the work in §3.

The main results of the present work are discussed in §4 and §5. In §4 we have reviewed a reconstruction procedure for the transmission coefficients and basic reproduction number 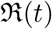. This procedure does not depend on the choice of any parameter and can be easily generalized for other similar models also. We have presented the best-fit values of the parameters with their 90% CI, §5 in the form a set of tables. We have presented the values of the parameters in the following two forms:

1. Model based
2. Country based

All the fitting parameters for the models being considered are summarized in Table (5) and full lists of parameters for different countries with different models is given at [Pra20a]. Here we only give the values of the effective reproduction for the countries (see Table (2) on the last date for which we have the data in Table (6).

The work we presented here assumes that spreading of a pandemic like Covid-19 happens homogeneously in space and time, however, we know that it is far from true. As the experience [ea03b] shows that “super-spread” events (SSEs) or rare events where, one particular infectious person interacts with a very large number of susceptible people over a short period of time have the maximum impact. In these situation the average measures like 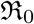 are not very informative. In the present work we data for a set of countries to constrain the parameters of the SIR and SIRD model one similar exercise with SIRD model for India is done in [ea20q].

In the present work we have considered only time variation of *β*, however, from the data we can see that other transmission coefficients such as *γ* and *δ* also change with time, although not that much, mainly because they depend more on the nature of the disease and less on the mitigation and other social measures. Significant change in the values of *γ* and *δ* can take place only due to medical interventions.

**Figure 15:**
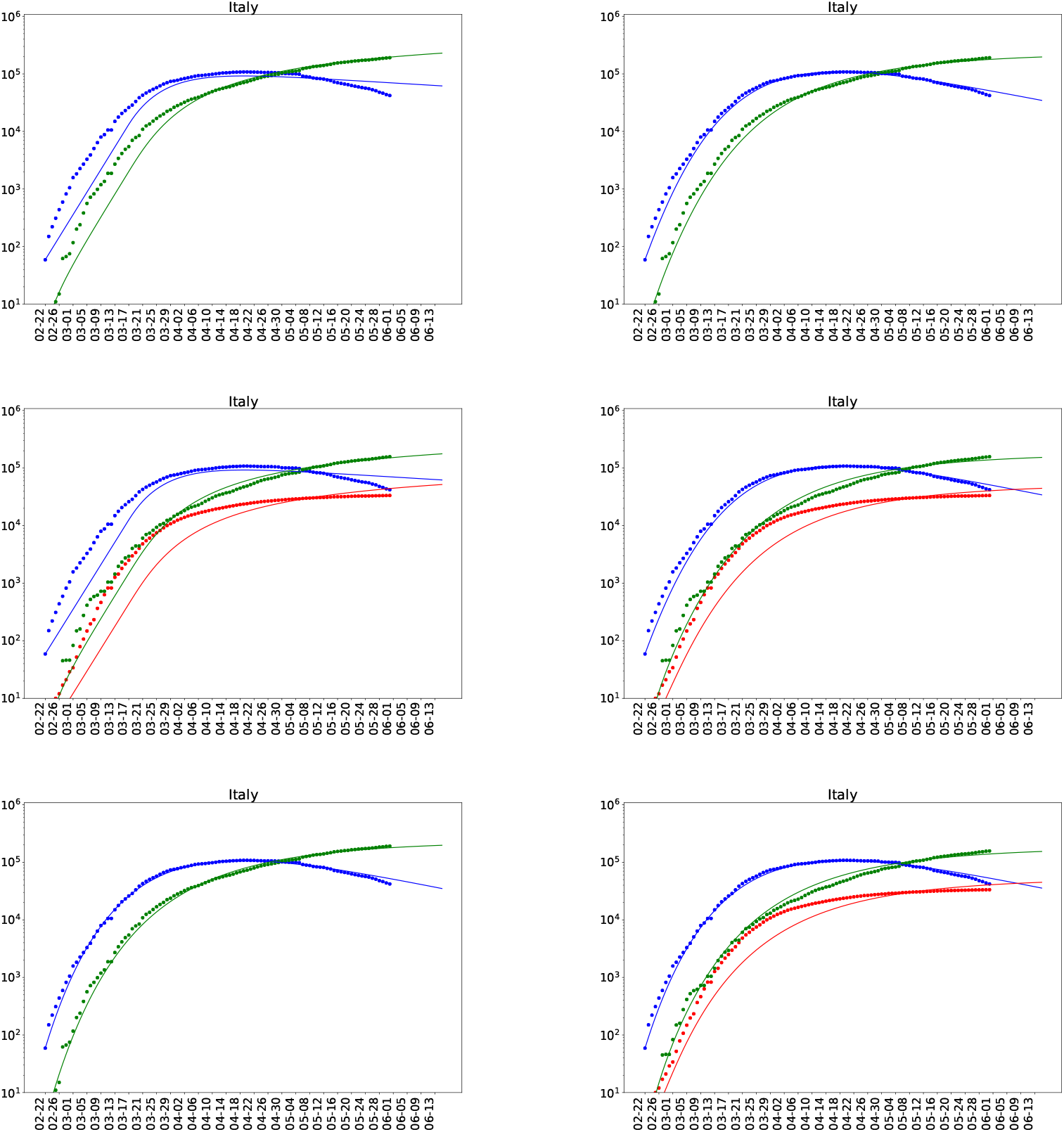
Fitting models with data for Italy. The panel from the top left to the bottom right are for the Models (1) to Model (6) respectively.

## Data Availability

All the data as well the code used in the work is available from the GitHub page of the author.

https://github.com/jayanti-prasad/PyCov19

## Acknowledgment

The author would like to thank Dr. Gaurav Goswami for comments and feedback. At present the author works as an independent researcher and data scientist and the work presented here is not supported by any public or private agency. The author will be thankful to any agency, individual or individuals who come forward to sponsor/support this and other similar works on Covid-19.

